# Integrating genomic variants and developmental milestones to predict cognitive and adaptive outcomes in autistic children

**DOI:** 10.1101/2024.07.31.24311250

**Authors:** Vincent-Raphaël Bourque, Zoe Schmilovich, Guillaume Huguet, Jade England, Adeniran Okewole, Cécile Poulain, Thomas Renne, Martineau Jean-Louis, Zohra Saci, Xinhe Zhang, Thomas Rolland, Aurélie Labbé, Jacob Vorstman, Guy A. Rouleau, Simon Baron-Cohen, Laurent Mottron, Richard A. I. Bethlehem, Varun Warrier, Sébastien Jacquemont

## Abstract

Although the first signs of autism are often observed as early as 18-36 months of age, there is a broad uncertainty regarding future development, and clinicians lack predictive tools to identify those who will later be diagnosed with co-occurring intellectual disability (ID). Here, we developed predictive models of ID in autistic children (n=5,633 from three cohorts), integrating different classes of genetic variants alongside developmental milestones. The integrated model yielded an AUC ROC=0.65, with this predictive performance cross-validated and generalised across cohorts. Positive predictive values reached up to 55%, accurately identifying 10% of ID cases. The ability to stratify the probabilities of ID using genetic variants was up to twofold greater in individuals with delayed milestones compared to those with typical development. These findings underscore the potential of models in neurodevelopmental medicine that integrate genomics and clinical observations to predict outcomes and target interventions.

## INTRODUCTION

Parents of children diagnosed with autism in early childhood invariably question their child’s future cognitive and adaptive development: will they be able to communicate, interact socially, and live independently?^1^

Early signs of autism, including differences in social communication and repetitive or restricted behaviour, often manifest around 12 to 18 months (figure 1A).^2,3^ These first concerns are also often preceded or accompanied by delays in language development, and in a more limited proportion of individuals, by delays in motor milestones.^4^ Formal diagnoses of autism can be established as early as 18 months (with 93% diagnostic stability^5^), but in most settings the earliest age of diagnosis is around 3-4 years of age.^2^ In toddlers and children with a suspected or confirmed diagnosis of autism, there is a broad uncertainty regarding future development and needs. Autistic individuals can display a broad diversity of strengths^6^ and disabilities, which might not directly correspond to their early developmental presentation.^7^

**Figure 1.**
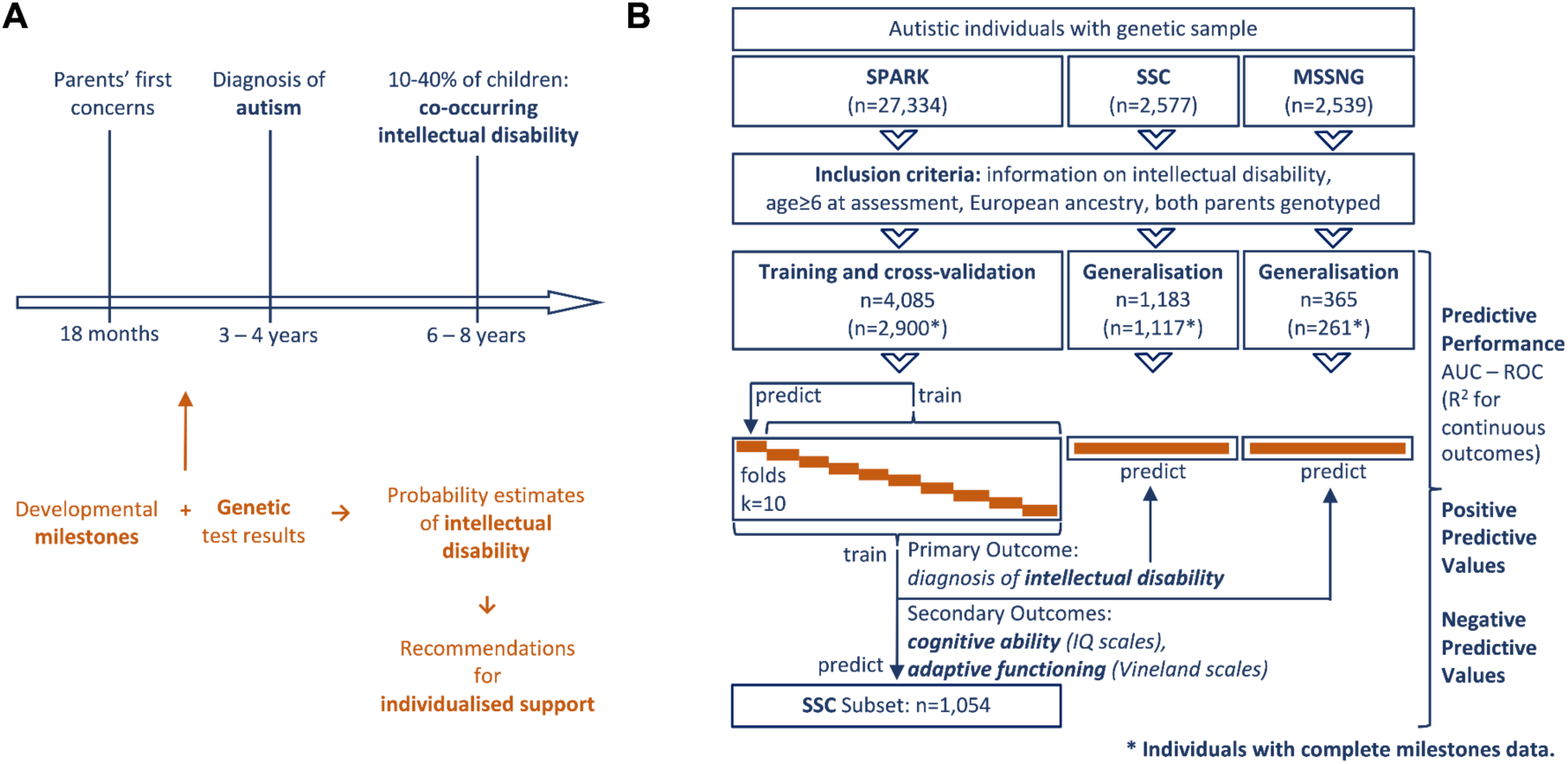
Schematic overview of the study. (A) Study rationale: There is often a long period of uncertainty between parents’ initial concerns, an autism diagnosis, and the identification of co-occurring ID in 10-40% of autistic children. Genetic testing is frequently conducted at or before the diagnosis of autism. Therefore, predictive models that integrate genetic testing results with developmental milestones could help clinicians provide parents with accurate information about their child’s expected developmental trajectory such as to offer the child appropriate support. B) Study flowchart: We trained models to predict ID in the SPARK cohort, estimating their out-of-sample predictive performance using cross-validation. Additionally, we evaluated the generalisability of these models in the SSC and MSSNG cohorts. AUC ROC: Area under the ROC curve. IQ: intellectual quotient.

Among all diagnoses recorded in the paediatric setting, a diagnosis of intellectual disability (herein, ID) may be one of the most predictive of future adaptive skills in adulthood.^8–10^ ID, defined by impairments in both cognitive ability and adaptive functioning, occurs in approximately 10-40% of autistic individuals,^11,12^ and can be diagnosed with relative confidence after the age of 6-8 years.^13^

Accurately predicting and anticipating ID would be useful, because with tailored support, individuals with ID can engage in significant relationships within and beyond their families (housemates, group activities, co-workers), develop independence in some daily-living skills, and engage in meaningful programs and recreational activities, supported employment, and volunteering positions.^14^ Clinicians currently use screening methods such as the Ages and Stages Questionnaire and the Bayley Scales of Infant Development, for the early identification of children who will later develop ID. The predictive ability of these tools remains, however, low, especially before 24 months of age.^15,16^ The current "wait-and-see" approach^17^ often overlooks a child’s specific strengths and challenges until there is a significant mismatch^18,19^ between these, the environmental demands, and the provided support.^20^ This mismatch can increase as a child grows older, resulting in higher levels of stress, school failure, and social misunderstandings.

Some autistic children, especially those diagnosed in early childhood or with other co-occurring conditions, undergo genetic testing.^21^ Additionally, in a growing number of individuals, genetic testing conducted for medical concerns during the neonatal period detects variants that are associated with neurodevelopmental conditions^22^. While the evidence for an association with autism and/or ID is established for an increasing number of genetic variants^23,24^, their effect size or penetrance has often not been accurately quantified.

The current goal of precision medicine introduces a new challenge: providing relevant predictions at the individual level.^25^ Recent genetic research exemplifies how common and rare genetic variants - including copy number variants (CNVs), and single nucleotide gene-disrupting variants - are associated with different adaptive and cognitive profiles in autistic individuals.^26–28^ However, highly significant statistical associations and large effect sizes derived from group-level comparisons do not necessarily translate into clinically relevant individual-level predictions.^29^ Consequently, there is a need for models to assist clinicians in accurately expressing the level of uncertainty in outcome predictions based on these genetic variants. This is critical in the field of autism and neurodevelopmental conditions broadly, considering that most genes with robust association with autism and ID are not fully penetrant, and their effect-size on neurodevelopmental phenotypes in population-based samples are systematically lower than previously estimated based on clinically referred cohorts.^27,30,31^

Recent data-driven approaches integrating clinical and genetic data into probability models have been developed in other medical fields such as oncology or cardiovascular health.^32,33^ This has resulted in improved predictive performance with genetic information complementing rather than replacing conventional predictors.^34–38^ This raises the question: to what extent can a similar approach be implemented in clinical practice for neurodevelopmental conditions? Currently, the identification of children who may require additional support due to developmental disabilities relies on the clinician’s experience. Increasingly, clinicians intuitively attempt to integrate information from genetic findings in their assessment, but thus far without the assistance from any predictive models as used in other medical fields.

In this study, we aimed to develop and validate models integrating genetic variants and early developmental milestones to predict the probability of developing ID in toddlers and children receiving a diagnosis of autism. Such models could allow clinicians to accurately communicate to families both the predictive informativeness and the level of uncertainty associated with interpreting genetic test results in the context of autism.

We asked: (1) Does integrating different classes of genetic variants together improve their performance for predicting ID? (2) Does the combination of genetic variants with developmental milestones improve predictive performance compared to milestones alone? (3) Do genetic variants and developmental milestones predict different cognitive and adaptive dimensions?

To do so, we analysed data from 5,633 autistic participants from three cohorts (figure 1B). We predicted probabilities of ID based on age at attaining five early developmental milestones along with language regression, polygenic scores, and four classes of rare genetic variants. To assess the potential for use in varied clinical settings, we tested out-of-sample performance, that is, the ability to provide accurate predictions for newly ascertained individuals. This was done using cross-validation in the SPARK cohort, and the models’ generalisability was further tested in SSC and MSSNG cohorts (see Inset: Definition of key technical terms).

Note on terminology: We chose to use the preferred term^39^ “autistic individual”; and “probability” or “likelihood” of ID, which do not have the connotation of danger or harm that the term “risk” could convey.

**Out-of-sample predictive performance:** the ability of a model - developed using data from one sample of individuals - to accurately predict an outcome (in this study, intellectual disability) in newly ascertained individuals, based on their clinical characteristics (e.g. developmental) and/or laboratory testing (e.g. genetic).

**Cross-validation:** a technique for estimating a model’s out-of-sample predictive performance within a single sample, by randomly dividing that sample into multiple groups of individuals and repeatedly training and evaluating the model’s performance on these distinct groups.

**Generalisability:** a model’s out-of-sample predictive performance in an entirely new sample, which might consist of individuals ascertained differently and in another clinical centre.

**Area under the Receiver Operating Characteristic curve (AUC ROC):** a gold-standard measure of a model’s global performance in providing probability estimates that accurately distinguish individuals that will develop an outcome from those that will not.

**Positive Predictive Value (PPV):** the proportion of individuals who develop an outcome, among all those predicted by the model to develop this outcome.

**Negative Predictive Value (NPV):** the proportion of individuals who do not develop an outcome, among all those predicted by the model to not develop this outcome.

**Sensitivity**: the proportion of individuals that were correctly identified by the model, among all those who develop an outcome. Also known as the true positive rate.

**Specificity**: the proportion of individuals that were correctly identified by the model, among all those who actually do not develop an outcome. Also known as the true negative rate.

**Probability stratification:** the ability to provide a range of probability estimates of an outcome according to each individual’s clinical characteristics (e.g. developmental) and/or laboratory testing (e.g. genetic).

**Inset: Definition of key technical terms.**

## RESULTS

### 1. Descriptive statistics of the cohorts

A total of 5,633 autistic individuals were included across three cohorts (inclusion criteria in figure 1A). The characteristics of the participants are detailed in Table Participants were diagnosed with autism at a median age of 4 years, even though their parents reported concerns starting from 18 months of age. ID occurred in 13.7 to 22.0% of individuals, depending on the sample, and was assessed or reported at a median age of 10 to 11 years old. Although, on average, a later diagnosis of ID allows for greater diagnostic stability^5^, this may be too late to provide the appropriate support for co-occurring ID. Consequently, an earlier prediction of co-occurring ID may be useful.

**Table 1.** Participant characteristics. Autistic individuals were included from three distinct cohorts: SPARK, Simons Simplex Collection (SSC), and MSSNG. For each milestone, we present the median [interquartile range, IQR], and the percentage delayed as compared to the 90th general population percentile (Perc.). Constrained genes are intolerant to mutations and therefore likely to have deleterious impacts on development^41^. Developmental Disorder (DD) genes represent a smaller set of genes previously associated with severe developmental disorders.^24^ Delayed milestones were defined based on the 90^th^ general population percentile.^42^

|  |  | Cohort used for training and cross-validation | Cohorts used for generalisation |  |
| --- | --- | --- | --- | --- |
|  |  | SPARK<br>(n=4,085) | SSC<br>(n=1,183) | MSSNG<br>(n=365) |
| Profile of participants | Sex, Female (%) | 20.2% | 12.8% | 22.5% |
|  | Age at parents' first concern, median [interquartile range, IQR], months | 18.0 [12.0, 30.0] | 18.0 [14.0, 30.0] | 18.0 [12.0, 30.0] |
|  | Age at diagnosis of autism, median [IQR], years | 4.25 [2.83, 7.00] | - | - |
|  | Age at ID assessment, median [IQR], years | 11.3 [8.42, 14.9] | 9.67 [7.92, 12.8] | 10.1 [8.21, 12.9] |
| Outcome | Diagnosis of ID (%) | 20.8% | 22.0% | 13.7% |
| Developmental predictors, median [IQR] (% delayed) | age at independent walking (general population 90<sup>th</sup> perc: 18 months) | 13.0 [12.0, 15.0]<br>(delay in 16.7%) | 12.0 [11.0, 15.0]<br>(delay in 10.4%) | 13.0 [12.0, 16.0]<br>(delay in 16.8%) |
|  | age at first word (general population 90<sup>th</sup> perc.: 15 months) | 14.0 [11.0, 24.0]<br>(delay in 48.6%) | 18.0 [12.0, 30.0]<br>(delay in 65.4%) | 18.0 [12.0, 36.0]<br>(delay in 66.9%) |
|  | age at first phrase (general population 90<sup>th</sup> perc.: 24 months) | 24.0 [18.0, 42.0]<br>(delay in 60.3%) | 36.0 [24.0, 48.0]<br>(delay in 78.5%) | 36.0 [24.0, 48.0]<br>(delay in 77.4%) |
|  | language regression (occurs on average at 21.8 months in autism)<sup>40</sup> | 22.7% | 15.4% | 16.6% |
|  | bladder control (months) | 42.0 [36.0, 54.0] | 43.0 [36.0, 54.0] | 45.5 [36.0, 54.0] |
|  | bowel control (months) | 48.0 [36.0, 60.0] | 48.0 [37.0, 60.0] | 48.0 [36.0, 60.0] |
|  |  | impacting constrained genes \| impacting DD genes<br>(2,971 genes \| 285 genes, genome-wide) |  |  |
| Carriers of genetic variants | deletions | 3.3% \| 0.8% | 2.3% \| 0.4% | 1.1% \| 0.5% |
|  | duplications | 4.8% \| 0.7% | 4.4% \| 0.6% | 4.7% \| 1.1% |
| | *de novo* missense variants with MPC≥2 | 3.0% \| 1.5% | 4.1% \| 2.1% | 3.3% \| 1.6% |
|  | *de novo* loss-of-function (LoF) variants | 7.1% \| 3.4% | 11.4% \| 3.9% | 7.4% \| 2.5% |
|  | any of the aforementioned variants | 17.3% \| 6.2% | 20.6% \| 6.9% | 16.2% \| 5.8% |

The ages at attaining developmental milestones were used as predictors in the analyses. We observed patterns typical of autism samples^4^: word delay in 48.6 to 66.9% (i.e. >90th general population percentile^42^) and phrase delay in 60.3 to 78.5% of individuals; walking delay occurred in a smaller portion, 10.4 to 16.8% of individuals, and language regression in 15.4 to 22.7% of individuals, similar to previous retrospective reports.^40^

In addition to polygenic scores (PGS) for cognitive ability^43^ and autism^44^, we ascertained copy number variants (i.e. deletions and duplications) and sequence variants (i.e. *de novo* loss-of-function - LoF - and missense variants). We observed that 16.2 to 20.6% of autistic individuals were carriers of at least one of those classes of variants that disrupted any of 2,971 constrained coding genes genome wide. Constrained genes are relatively intolerant to mutations and therefore more likely to have deleterious impacts on development. These genes were previously identified^41^ based on a lower observed than expected (LOEUF<0.35) number of disrupting variants in large samples. In addition, we ascertained the same classes of variants disrupting a set of 285 genes previously associated with severe developmental disorders (DD).^24^ The latter were identified in 5.8 to 6.9% of autistic individuals.

### 2. Predictive value of combining different classes of genetic variants Evaluating genetic variants with ROC curve analysis

We first investigated if genetic variants alone could predict ID among autistic individuals (figure 2A). In the SPARK cohort, all genetic models were statistically significant (likelihood ratio test, p=1.1x10^-17^ for the model including all classes of variants) and we observed an increase in cross-validated predictive performance measured with the AUC ROC with sequentially incorporating the different classes of common and rare genetic variants into the model. The predictive performance of the model combining cognitive ability PGS and autism PGS resulted in an area under the ROC curve (AUC ROC) of 0.529 (95%CI: 0.512, 0.545). With the inclusion of constrained and DD gene deletions and duplications this was 0.532 (95%CI: 0.515, 0.548), and when also incorporating de novo LoF and missense variants, 0.565 (95%CI: 0.545, 0.586).

**Figure 2.**
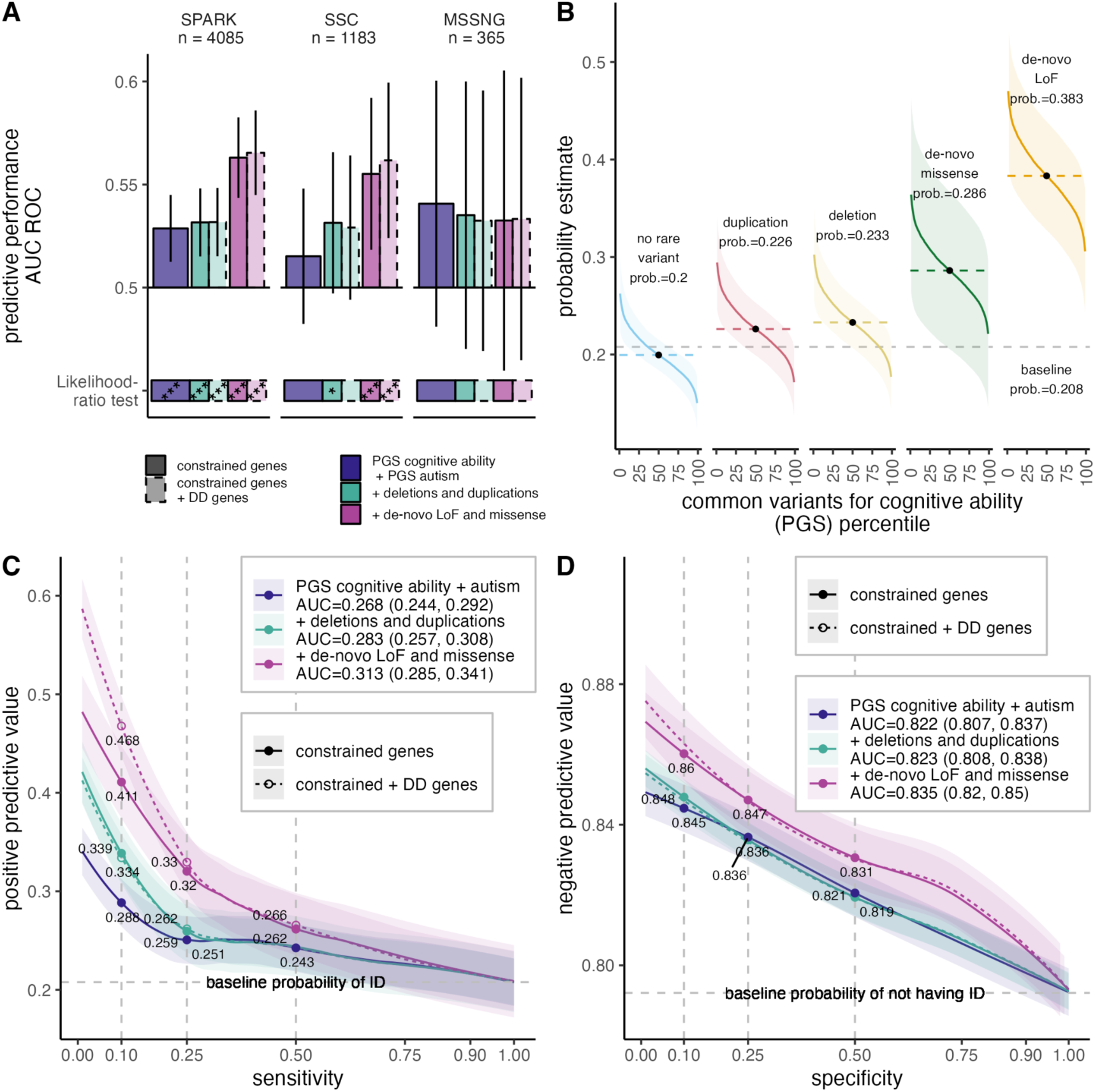
Genetic analyses to predict intellectual disability in autistic individuals. (A) Predictive performance of combining different classes of genetic variants, assessed using cross-validation in the SPARK cohort and evaluated for generalisability in the SSC and MSSNG cohorts. The predictive performance of each model is tested using two complementary methods: the likelihood-ratio test, and the AUC ROC (Area Under the ROC Curve, with its 95% confidence interval). Genetic variants were included sequentially into the models: polygenic scores (PGS) for cognitive ability and autism, deletions and duplications, de novo loss-of-function (LoF) and missense variants. Rare variants were annotated based on constrained genes alone, or combined with developmental disorder (DD) genes. Benjamini-Yekutieli adjustment for multiple comparisons, *p<0.05, **p<0.01, ***p<0.001. (B) Probability estimates of ID for carriers of rare variants in constrained genes are shown (coloured curve) as a function of their common variant background, measured by polygenic score (PGS) for cognitive ability. Shaded areas representing 95% confidence intervals. Dashed lines indicate the average probability of ID for carriers of each class of rare variant, as well as for individuals who do not carry any rare variant. The baseline probability corresponds to the prevalence of ID in the total sample, irrespective of genetic variants. LoF: loss-of-function. (C) PPVs achieved at different probability thresholds reflect the trade-off with the model’s sensitivity. The horizontal grey dashed line represents the baseline prevalence of ID in the sample. Annotations show that for a given sensitivity value (displayed for 10%, 25%, and 50% sensitivities), models integrating different classes of genomic variants yield progressively higher PPVs. Shaded areas represent 95% confidence intervals. The Area Under the Curve (AUC) reflects the overall ability to predict ID and should be compared to the sample’s baseline ID prevalence (20.8%). (D) As a counterpart to panel C, NPVs achieved at different probability thresholds reflect a trade-off with the model’s specificity. The AUCs reflect the overall ability to predict the absence of ID, and should be compared to the sample’s baseline prevalence of not having ID (79.2%).

#### Generalisation to different cohorts

We tested whether the model’s predictive performance was generalisable to other cohorts with different ascertainment (figure 2A). The model trained on the SPARK cohort demonstrated similar predictive performance when used to predict in the SSC cohort (AUC=0.562; 95%CI: 0.524, 0.599; likelihood-ratio test p=1.2x10^-6^), and in the smaller MSSNG cohort (AUC=0.533; 95%CI: 0.465, 0.602; likelihood-ratio test p=0.41).

#### Examining model predictions

We examined the probability estimates (Figure 2B) obtained from combining PGS for cognitive ability with rare genetic variants of different classes. The estimated probability in individuals with a bottom-decile PGS (i.e. implying a high-liability for ID) was comparable to the probability in carriers of deletions or duplications of a constrained gene. In individuals with a higher probability due to a de novo LoF variant, the top and bottom decile PGS were associated with predicted probabilities of ID of <34.0% and >43.0%, respectively, as compared to <17.1% and >23.2% in those without any rare variant. In other words, combining rare and common variants allowed further stratification and improved discrimination between low versus high probabilities of ID.

#### Evaluating positive and negative predictive values

To evaluate the clinical utility of this model, we examined trade-offs between positive predictive values (PPVs) and sensitivity, as well as between negative predictive values (NPVs) and specificity (see Inset: Definition of key technical terms), across the full range of combinations of genetic variants (figure 2C and D).

We observed that the addition of each class of variant into the model (figure 2C) consistently improved PPVs. Certain combinations of variants reached a PPV of 46.8%. In other words, 46.8% of the individuals predicted to have ID by the model had a diagnosis of ID (twofold increase compared to the baseline prevalence of 20.8%). Such high-liability combinations of variants occurred in a small proportion of individuals, thus correctly identifying 10% (sensitivity) of individuals who will develop ID.

Gains in NPVs (figure 2D) were more modest. The NPVs reached 86.0%, as observed at the lowest 10% of specificity, compared to the 79.2% baseline prevalence of not having ID. This suggests that genetic variants, when used in isolation, are more effective at predicting the presence of ID (figure 2C) than excluding it (figure 2D).

Many variants (with mild-to-moderate effect sizes) integrated in our model would either not be reported by most diagnostic laboratories, or be reported as “variants of unknown significance”. Therefore, we compared our model’s PPVs with that of variants that would currently be reported to clinicians as “pathogenic” by diagnostic laboratories. The carrier status of deletions or *de novo* LoF variants disrupting DD-associated genes^24^ had a PPV of 49.8%, similar to that of the model integrating all classes of variants, albeit with a 3-fold smaller sensitivity (3.3%, Supplementary Table 6). This indicates that models integrating different classes of genetic variants achieve PPVs similar to those of currently reported pathogenic variants in a threefold larger group of individuals (i.e. improved sensitivity).

#### Sensitivity analyses

We assessed whether other factors may have hampered the models’ performance in predicting ID. We evaluated the impact of removing individuals who had prenatal exposure to alcohol or drugs, oxygen supplementation at birth, intraventricular haemorrhage, meningitis, or encephalitis (6.1% of individuals in SPARK). Additionally, because cognitive assessments of non-speaking individuals may not accurately reflect their cognitive abilities, we tested the impact of removing non-speaking individuals (10.8% of individuals in SPARK). The models’ performance for predicting ID based on genetic variants when these individuals were removed was similar as to when they were included (Supplementary Table 4). Also, to determine if including interactions and potential nonlinear relationships would improve predictions, we trained a Random Forest algorithm, which resulted in performance comparable to the logistic model (Supplementary Table 2).

### 3. Integrating genetic variants and developmental milestones to predict outcome

When a child is referred for atypical development, clinicians commonly take into account developmental history to assess the probability of developing ID, based on normative data and their clinical experience. We aimed to evaluate the predictive informativeness of genetic test results on the background of this clinical information.

First, we tested the predictive value of developmental history, by sequentially including in a model the age of attaining 5 milestones, as well as the presence of language regression, in the order these typically occur (figure 3A, lower panel).^42^ As expected, the predictive performance increased as we added developmental milestones that are achieved at older ages, closer to the diagnosis of ID. The ages of bowel and bladder training provided little additional predictive information.

**Figure 3.**
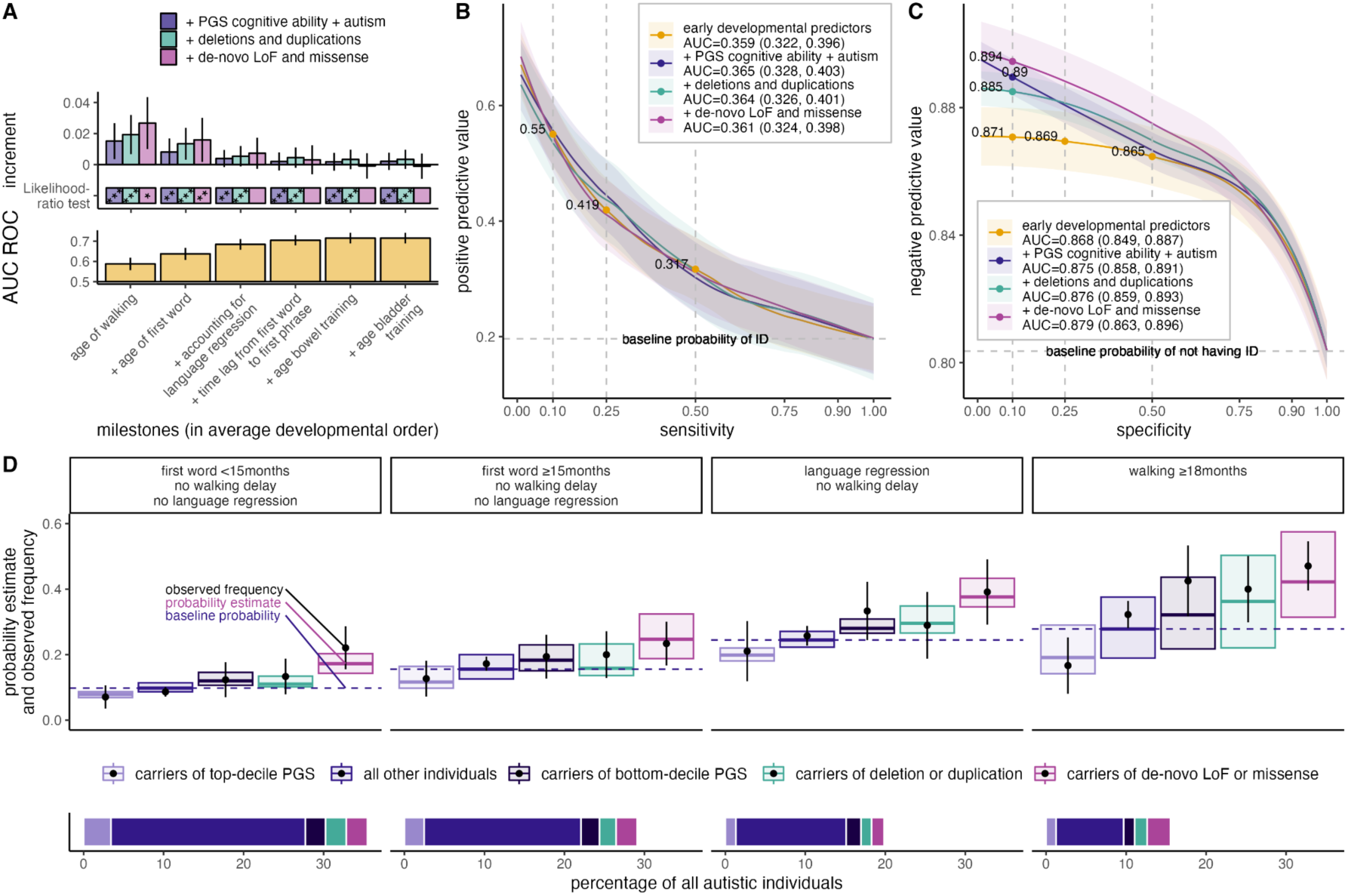
Combining genetic variants and developmental milestones to predict intellectual disability in autistic individuals. (A) Predictive performance: The lower panel shows the AUC ROC of developmental models with sequentially adding milestones in the typical order of child development. The upper panel shows to what extent genetic variants provide additional information when combined with early milestones. The predictive performance of each model is compared against the model consisting of the developmental milestones using two methods: the likelihood-ratio test, and the difference in AUC ROC (with its 95% confidence interval). Genetic variants were included sequentially into the models: polygenic scores (PGS) for cognitive ability and autism, deletions and duplications, de novo loss-of-function (LoF) and missense variants. Benjamini-Yekutieli adjustment for multiple comparisons, *p<0.05, **p<0.01, ***p<0.001. (B) PPVs achieved at different probability thresholds reflect a trade-off with the model’s sensitivity. The models’ PPVs should be compared against the horizontal grey dashed line, which represents the baseline prevalence of ID in the sample. Annotations show that at a given values of sensitivity (displayed for 10%, 25%, and 50% sensitivities), the PPVs are similar for models that include milestones plus genetic variants compared to that including only milestones. Shaded areas represent 95% confidence intervals. The Area under these PPV-Sensitivity curves (AUC) reflect the overall ability to predict ID, compared to the sample’s baseline ID prevalence. (C) As a counterpart to panel B, NPVs achieved at different probability thresholds reflect a trade-off with the model’s specificity. Annotations show that with keeping specificity values constant (here displayed for 10%, 25%, and 50% specificities), the NPVs are higher for the model including polygenic scores (PGS) and milestones, compared to the model including only milestones. The AUCs under the NPV-Specificity curves reflect the overall ability to predict the absence of ID, compared to the sample’s baseline prevalence of not having ID. (D) Probability of ID based on combinations of developmental milestones and genetic variants. Groups are defined based on common reasons for clinical referral. The box plot represents the distribution of out-of-sample model predictions (median and interquartile range). The black dots and 95% confidence intervals represent the observed frequencies of intellectual disability within the same group. The bottom panels indicate the percentage of autistic individuals assigned to each group. PGS: polygenic score for cognitive ability. LoF: loss of function.

Then, we assessed the added predictive value of genetics (figure 3A, upper panel) at each developmental stage, compared to milestones alone. The added predictive value of genetic variants was contingent on the performance of milestones, such that it decreased when milestones occurring at an older age were included in the model, suggesting that genetic variants contribute more prediction early in development when less information on milestones is available. We then focused on a model integrating the ages of walking and first words as these milestones are typically observed at the time of referral for autism assessment. The predictive performance was AUC ROC=0.637 (95%CI: 0.608, 0.667). The sequential additions of polygenic scores, copy number variants, and *de novo* coding variants into the model were respectively associated with increments in AUC ROC of 0.008 (likelihood-ratio test, p=1.0x10^-3^), 0.013 (p=1.6x10^-5^) and 0.016 (p=9.3x10^-3^). This complete model had an AUC ROC of 0.653 (95%CI: 0.625, 0.681).

#### Evaluating positive and negative predictive values

To understand the strengths and limitations of this model for clinical predictions, we examined PPVs and NPVs across combinations of genetic and developmental trajectories (figures 3B and C). We expected that incorporating genetic information alongside developmental milestones would primarily improve PPVs, as clinicians generally consider the carrier status of a genetic variant more clinically significant than its absence. However, we observed that the addition of PGS to developmental milestones improved NPVs (Figure 3C). Certain combinations of variants (or their absence) and milestones reached a NPV of 89%. In other words, 89% of the individuals whom the model predicted would not have intellectual disability (ID) indeed did not have a diagnosis of ID (which should be compared to the baseline prevalence of not having ID).

#### Stratification of probability estimates

While our models were trained using comprehensive developmental and genetic information, we aimed to assess their predictive accuracy in clinical situations where children are referred for specific concerns. To illustrate the models’ ability to differentiate between high and low probabilities of ID, we computed probability estimates within four groups. These groups were characterised by common reasons for referral to developmental clinics, such as delayed (>90^th^ percentile) motor or language milestones or language regression.^42^

We examined (figure 2D) how the consideration of genetic test results could further stratify the probabilities of ID within each group. The observed frequencies of ID for each group were concordant with the probabilities of ID predicted by the model. We observed a wide spectrum of probabilities resulting from the combination of genetic variants and developmental history. For example, the stratification between low versus high probabilities provided by genetic variants was increased twofold in individuals with significant developmental delays compared to those without such delays: for the group with the highest developmental probability estimate (walking delay), a top-decile protective PGS was associated with a 16.7% observed proportion of ID, whereas carrier status of a *de novo* missense variant with 47.1% proportion of ID. In comparison, in the group with the lowest developmental probability estimate (i.e. no delays and no regression), top-decile PGS was associated with a 7.1% proportion of ID, whereas carrier status of a de novo missense variant with 22.1% proportion of ID.

#### Comparison of higher-probability and lower-probability groups

To illustrate how varied combinations of genetic variants and milestones may result in high or low probabilities of ID, we compared developmental and genetic predictors in the top and bottom 5% of predicted ID probabilities (each n=145). Again, the observed and predicted frequencies of ID were concordant: in the top 5% of individuals with high probability combinations, these were respectively 47.6% and 54.4% (IQR: 47.2, 64.6); and in the bottom 5% of individuals, respectively 7.6% and 7.29% (IQR: 6.78, 7.70).

Individuals in the top 5% probability group achieved milestones later, with walking at median 24.0 (IQR: 18.0, 30.0) and first word 42.0 (IQR: 27.0, 60.0) months; language regression was frequent (35.2%). More than half carried rare variants in constrained genes (54.5%) among which 27.6% in DD genes. Their PGS for cognitive ability and autism were below average, -0.319 (IQR: -0.936, 0.399) and -0.184 (IQR: -0.958, 0.414), respectively.

Conversely, individuals in the bottom 5% probability group achieved milestones earlier, with walking at median 10.0 (IQR: 9.00, 12.0) and first word 9.00 (IQR: 6.00, 11.0) months; none had language regression. Rare variants in constrained genes were seldom observed (2.8%; 0.7% of which in DD genes), and their PGS for cognitive ability and autism were above average, 1.03 (IQR: 0.362, 1.73) and 0.0422 (IQR: -0.691, 0.886), respectively.

#### Out-of-sample generalisation

The predictive performances provided by the combination of developmental milestones and genetic variants were generalised SSC and MSSNG (Supplementary Table 3).

### 4. Differential prediction of cognitive and adaptive dimensions

We asked whether some of the limitations of predictive performance may be explained by the inconsistent relationship between cognitive ability and adaptive functioning observed in autism.^45,46^ Impairments in both these dimensions are required to meet criteria for ID in both DSM-5 and ICD-11. Congruent with previous reports, we observed that a large proportion of high functioning (i.e. with IQ>70) autistic individuals had more impairment in adaptive functioning than expected based on their intellectual quotient (IQ, figure 4A), as reflected by the nonlinear relationship between IQ and Vineland scales. This was observed across all adaptive subscales, with the weakest relationship observed between motor skills and verbal IQ (r=0.32).

**Figure 4.**
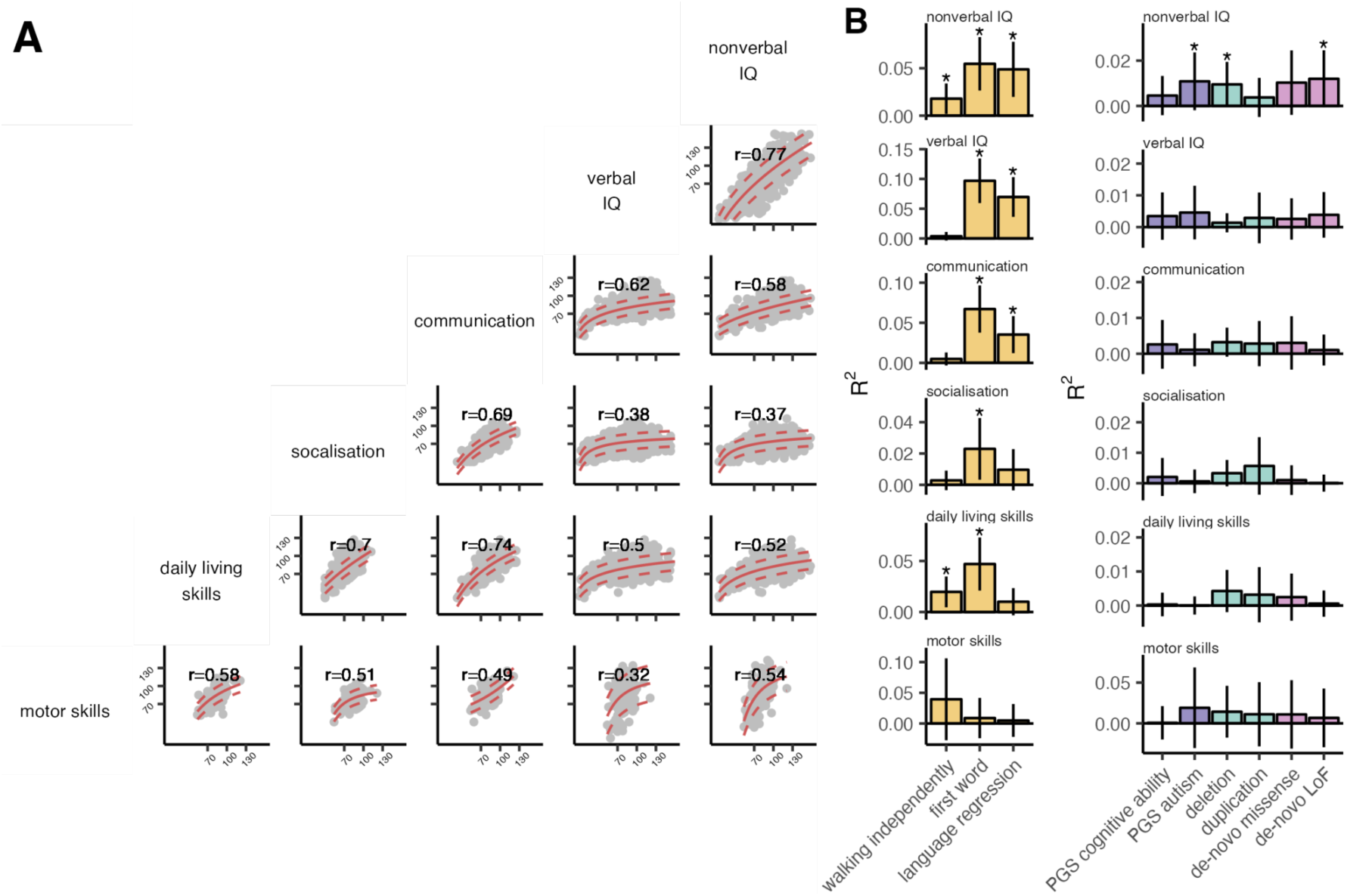
Differential prediction of adaptive and cognitive dimensions using genetic variants and developmental milestones. (A) Correlations between cognitive and adaptive dimensions, measured by IQ and Vineland adaptive behaviour scales respectively. The Spearman correlation matrix indicates non-linear relationships ranging from r=0.32 to 0.77. Red lines represent the median, 10th and 90th percentiles of the non-linear trend computed using GAMLSS. X and Y-axis values are standardised scores with mean 100, and standard deviation 15. (B) Predictive performance for cognitive and adaptive dimensions. The predictive performance of developmental and genetic predictors varies across cognitive and adaptive dimensions, as shown by their distinct patterns of explained variance (R²). Significance is evaluated against the null model, *p<0.05 (Benjamini-Yekutieli adjustment for multiple comparisons). All subscales and predictors were evaluated in the same sample of 1,054 participants, except for the optional subscale or motor skills, which was assessed in a subset of 160 participants where motor impairments were suspected by examiners.

Therefore, we asked whether genetic variants and early milestones were more predictive of some defining features of ID rather than others: verbal and non-verbal IQ, as well as the four Vineland subscales of adaptive functioning (figure 4B, Supplementary Table 5). We evaluated this in a subset of autistic individuals with available data from the SSC sample. Significant milestones explained on average four times more variance than genetics, with on average 4.8% of variance explained by milestones versus 1.08% for the genomic variants assessed in this study (including significant effects only). As expected, language milestones were more predictive of verbal IQ. All classes of rare and common variants, as well as age of walking, were more predictive of nonverbal compared to verbal IQ. This observation is noteworthy, as cognitive assessments predominantly emphasising verbal aspects have been shown to overlook important strengths of autistic individuals.^47^

## DISCUSSION

Leveraging previous genetic studies that have associated rare and common genetic variants with ID and autism, we have trained and evaluated models to estimate the probability of ID in autistic individuals from three independent samples. These models integrate the broad spectrum of combinations of genetic variants and developmental milestones observed in autistic individuals and provide positive predictive values (PPVs) of up to 55%, accurately identifying 10% of ID cases. Combinations of variants that are typically not considered clinically relevant on their own by diagnostic laboratories can yield clinically significant PPVs. Notably, the ability to stratify the probabilities of ID using genetic variants was up to twofold greater in individuals with delayed milestones compared to those with typical development. This approach addresses a core ambition of precision medicine, which is the early identification of individuals who may benefit from targeted interventions.^25^

Currently, many national guidelines recommend screening of all children at 18 months of age for delays in development as well as autism.^48^ However, the predictive performance of current screening tools relying solely on early developmental milestones is limited, especially before 2 years of age.^15^ This emphasises the need for integrating additional sources of predictive information, such as results from genetic testing.

With the implementation of whole genome sequencing in many clinical settings, common variants are detected, but remain underutilised. Our results show that clinically significant predictions can be obtained by integrating common variants, as captured by polygenic scores (PGS), with various classes of rare genetic variants. This allows to explain, in part, what is described as variable expressivity in individuals carrying rare variants with large effect sizes. This approach builds on previous studies showing that the effects of rare variants are modulated by an individual’s genetic background of common variants.^26,32,49,50^. Previously, this was applied to a specific rare variant, the 22q11.2 deletion, where it was shown that the predictive power of PGS for ID was higher in carriers (due to an elevated baseline prevalence) than in the general population.^32^ The PPVs reached 63% for 22q11.2 deletion carriers with a top decile PGS, compared to the baseline prevalence of 41% among all carriers.

Here, we extend this approach to all autistic individuals. Including multiple combinations of rare and common variants - without any phenotypic information - the models attain PPVs of 46.8% compared to a baseline probability of 20.8% in this sample, and correctly identify 10% of individuals who will develop ID. Although a sensitivity of 10% is modest, we show that it represents a threefold improvement over an approach that focuses solely on the carrier status of developmental disorder-associated variants, as typically reported by diagnostic laboratories. PGS contribute to a large part of the predictive ability for ID, which may be partly because they are measurable in all individuals, highlighting their potential in clinical settings, especially when assessed alongside protein-disrupting variants.

Probability prediction in other medical fields such as cardiovascular health relies on quantitative tools that integrate multiple variables, each contributing modestly to predictive power, but collectively achieving clinically significant results.^51^ We expect that improving the prediction of neurodevelopmental outcomes will similarly require the integration of multiple developmental and genetic variables, each contributing incrementally to predictive performance. While the heritability of ID^52^ is high, a considerable portion of genetic variants contributing to ID remains unidentified to this day. Current polygenic scores explain ∼5% of variance in general cognitive ability^53^, which is low compared to other traits (with similar heritability) such as height where association studies have reached saturation and provide PGS explaining up to 40% of variance.^54^ Improved knowledge of the genetic variants involved in neurodevelopmental conditions, including GWAS performed across diverse ancestries, is expected to increase the predictive power of such models.

Any use of genetics to predict neurodevelopmental outcomes is impacted by pleiotropy, where a single genetic locus or variant influences multiple phenotypes. Most genetic variants disrupting constrained genes affect multiple developmental phenotypes, such as autism, ID^55^, as well as developmental milestones which, importantly, are not diagnostic criteria for autism nor ID.^56^ This pleiotropy may result from the expression of constrained genes across multiple cell types and tissues, leading to a wide range of associated phenotypes.^41^

As such, the pleiotropy of genetic variants across phenotypic predictors and outcomes represents an opportunity to improve predictive models by using developmental predictors as proxies for currently unmeasured genomic liabilities (e.g., unidentified gene-disrupting variants may be present in children with significantly delayed milestones). Consistent with previous findings that the developmental milestones of carriers of high-impact rare variants correlate with their cognitive and adaptive outcomes^57^, we observed that incorporating developmental milestones improved predictive performance compared to models based solely on genetic variants, highlighting the clinical relevance of continued developmental monitoring in carriers of these variants.

Conversely, we show that genetic variants bring additional predictive information compared to milestones alone. Specifically, the ability to discriminate between low versus high genetically predicted probabilities of developing ID was twice as high in children with an elevated baseline probability of ID due to delayed milestones; and incorporating genetic data, particularly PGS, with developmental milestones improved the identification of children with delayed milestones who did not develop ID.

### This study has several limitations

The retrospective evaluation of milestones may be susceptible to recall biases or the "telescoping effect”, wherein developmental events can be remembered by caregivers as more recent than they actually occurred.^58^ However, this mirrors current practices, where developmental history is gathered retrospectively at the time of an autism diagnosis.

Selection bias^59^ might lead to an over-representation of individuals without ID, including possibly those individuals who are carriers of variants in known autism/ID genes, since significant ID may exclude some individuals from an autism diagnosis and/or participation in research.^60^ This could reduce the predictive performance of models for predicting ID. Indeed, it was previously shown that in the SPARK cohort, more than 25% of autistic individuals carrying *de novo* LoF variants in a set of autism-associated genes had an IQ in the typical range.^27^

The predicted outcomes were limited to ID, and to cognitive and adaptive dimensions. Future research could investigate other outcomes for which prophylactic treatment or risk-alleviating measures exist, such as seizures or a psychotic episode^32^, in autistic individuals or in other groups that have an elevated baseline risk.

### Conclusion

This study highlights the feasibility of using predictive models to assist clinicians in their assessment of children referred for autism. Our findings show that the growing number of robust genetic associations with autism and ID cannot, in most cases, be used alone; but models combining all variants and developmental milestones may provide clinically relevant individual-level predictions. Such models can provide families with probability estimates for different developmental trajectories, along with the corresponding levels of uncertainty tied to the interpretation of developmental milestones and genetic findings in the context of autism.

## METHODS

### Sample selection

We included participants from 3 cohorts (figure 1B): the Simons Foundation Powering Autism Research (SPARK)^61^ version WES_1-2-3, Simons Simplex Collection (SSC)^62^, and MSSNG^63^ cohorts. Participants were included based on the following criteria: (1) Diagnosis: a professional diagnosis of autism spectrum disorder (herein referred to as autism) according to DSM-5-TR or corresponding categories from previous editions. This was based on caregiver report or self-report. (2) Genetic data: the genetic data from the proband and both parents had to meet quality control, detailed further below, as this is required for calling *de novo* variants, and (3) Outcome: documented information on ID, and age at least 6 years for greater reliability of this assessment.^64^ Analyses of developmental milestones were done in the subset of individuals for whom all milestones were available.

### Primary outcome

In the SPARK cohort, the occurrence of ID was determined based on a caregiver report of a professional diagnosis. In SSC and MSSNG cohorts ID was inferred from non-verbal IQ data using a threshold below 70. This method was chosen based on evidence suggesting that non-verbal assessments more accurately reflect the cognitive abilities of autistic individuals.^47^

### Secondary outcomes

In a subsample of the SSC cohort where data were consistently available, we assessed the subscales from the intellectual quotient (IQ), and the Vineland Adaptive Behaviour Scales.^65^

### Genetic quality control and imputation

Genomic analyses were obtained either by single nucleotide polymorphism (SNP) genotyping (Illumina 1Mv1, 1Mv3, or Omni2.5 arrays for SSC; Illumina Infinium Global Screening Array-24 for SPARK) or by Whole-Genome Sequencing (WGS: Complete Genomics or Illumina HiSeq 2000/X Ten for MSSNG). In the case of WGS, we used PLINK v1.9^66^ to convert the pVCF files into genotyping data format^67^. Using bcftools v1.13^68^ we removed indels, selected biallelic sites, and normalised the SNP dataset.

We performed quality control for each genetic sequencing technology separately using established criteria^69^ with PLINK v1.9.^66^ In brief, we removed individuals with genotyping rate <95%, excessive heterozygosity (±3 standard deviations from the mean), sample missingness >0.02, mismatch in reported and genetic sex, or families with Mendelian errors >5%. We removed SNPs with a call rate <98%, a minor allele frequency (MAF) <1%, deviation from Hardy-Weinberg Equilibrium (P <1×10^-6^), or >10% Mendelian error rate.

We assessed the population genetic structure using KING^70^. Considering that the current genome-wide association study (GWAS) summary statistics are primarily Euro-centric, we included individuals of comparable genetic background, i.e. with ≥85% probability of genetically inferred European ancestry.

We imputed the missing SNPs using the 1000 Genomes Project phase 3 (1KGP3) reference panel^70^ on the Sanger Imputation Server.^71^ We retained loci that were measured across all technologies. We merged the imputed files, converted them to PLINK files, and we removed SNPs with a poor imputation quality metric (INFO≥0.3), multiallelic (>2) variants, MAF <5%, call rate <98%, or significant deviation from Hardy-Weinberg Equilibrium (P <5×10^-7^).

To account for population stratification within the final sample, we computed the top 10 ancestry principal components (PCs) using the mds function in the KING population structure inference tool, with the 1000 Genomes Project data as a reference population.^70^

### Polygenic scores

To compute the PGS, we used GWAS summary statistics for autism and 11 traits genetically correlated to autism. These traits were previously selected from a total of 234 traits tested for genetic correlations with autism, as presented in the autism GWAS publication.^44^ We reasoned that given their correlation with autism, these genetic liabilities could potentially also be relevant with regards to the heterogeneity of outcomes among autistic individuals. For the autism GWAS statistics we included only the iPsych sample, such that there is no overlap between the base GWAS samples and the target samples of this study.

We computed PGS using polygenic likelihood scoring with continuous shrinkage (PRS-CS), which is one of the polygenic scoring strategies that yields the highest variance explained.^72^ To remove potential confounding effects of population structure, we residualised the top 10 ancestry principal components from each PGS within each cohort.

To select the most predictive combination among the 12 PGS, we used the technique of forward feature selection with 10-fold cross validation in the SPARK dataset, because these were partly collinear and varied significantly in their predictive performance for ID. The selection criterion was to optimise predictive performance assessed by the AUC ROC. This led us to retain for the analyses the two most predictive PGS: that for cognitive ability^43^ followed by that for autism^44^ (Supplementary Table 1), which we combined in an additive linear model as previously reported.^73^

### Copy number variants

We identified copy number variants (CNVs) from SNP arrays using two independent pipelines, PennCNV^74^ and QuantiSNP^75^. A consensus from these two pipelines was obtained using CNVision^76^, such as to minimise false discoveries. For MSSNG, we used read alignment data to compute CNV calling from WGS.^77^

We removed artefactual and mosaic CNVs using a previously published pipeline.^28,78,79^ We retained CNVs with call rate ≥95%, log R ratio standard deviation <0.35, B-allele frequency standard deviation <0.08, and |wave factor|<0.05. For consistency across technologies, we retained CNVs that overlapped ≥10 probes in each array technology.

CNVs were annotated with Gencode V19 (hg19) using ENSEMBL.^80^ Deletions and duplications were annotated with coding genes of which all isoforms were fully encompassed.

### *De novo* coding variants

We used previously published calls of *de novo* coding variants for SPARK, SSC^27^ and MSSNG^63^. We included all loss-of-function (LoF) variants, as well as missense variants with high variant-level Missense PolyPhen Constraint (MPC≥2).^56,81^

### Annotation of rare variants

We annotated CNVs and *de novo* coding variants with a gene-level constraint metric, the Loss-of-function Observed/Expected Upper bound Fraction (LOEUF) score.^41^ For all 4 classes of rare variants (deletions, duplications, *de novo* LoF, and missense), we employed a gene-based scoring strategy, assigning scores to each individual based on the count of rare variants, within each class, affecting constrained genes. Constrained genes were defined with LOEUF<0.35^30,41,78^, which represents n=2,971 constrained genes genome-wide. In addition, we evaluated a smaller set of n=285 genes^24^ potentially more specifically associated with Developmental Disorders (DD).

### Developmental milestones

Developmental milestones were assessed in all three cohorts across motor (walking), language (speaking words, phrases, language regression) and toileting (bladder and bowel control) domains based on retrospective reports by caregivers. Milestones were recorded as age of attainment in months, except for language regression which was binary-coded.

For SPARK, milestones were collected through the Background History Questionnaire filled by a caregiver or independent adult. For SSC and MSSNG, milestones were collected through the Autism Diagnostic Interview-Revised.^82^ The selected milestones are attained on average in the general population before 4 years of age^83^, and language regression occurs in a proportion of autistic individuals on average by 2 years of age^40^, thus, participants had aged past this period at time of assessment (≥6 years old), consequently this allows from complete records.

The ages at attaining the milestones were included as predictors within a multiple logistic regression. Previous research suggests that the relationship between the age at first word and IQ may be different in individuals with language regression^84^, therefore we included an interaction term between those two predictors. To include a maximum of information from the age at first phrase whilst also accounting for its collinearity (r=0.72) with the age at first word, we calculated the time lag between first word and first phrase, which is supported by previous accounts of a language acquisition plateau a proportion of autistic individuals.^84^

### Framework for model training and evaluation

Using 10-fold cross-validation in the SPARK cohort we trained the models and assessed their predictive performance on unseen data. In this approach, the dataset is divided into ten equal parts, or "folds," each with a stratified representation of the outcome variable. The model is trained on nine of these folds and tested on the tenth, iterating this process until each fold has served as the test set. This method, a standard in machine learning^85^, helps safeguard against overfitting by ensuring that the models maintain strong performance on unseen data^86^. By averaging the performance metrics across all folds, we obtain a robust estimate of the models’ out-of-sample predictive performance.^87^

To determine the individual and cumulative predictive contributions of each variable, we used multiple logistic regression, sequentially adding variables in a predetermined order:

A. Genetic models: We integrated polygenic scores (cognitive ability and autism), followed by CNVs (deletions and duplications) and *de novo* coding variants (LoF and missense variants). Sex was included as a covariate in all models, and we then adjusted the performance estimates to specifically reflect the contributions attributable solely to genetic variables.
B. Developmental models: Developmental milestones were incorporated sequentially based on the average age at which they are typically attained in child development^42^, and including sex as a covariate.
C. Integrated Models: Following the approach used in other medical fields for “integrated risk models”,^34,88^ we combined genetic variants in the same models with developmental milestones. We evaluated the increase in predictive performance resulting from the sequential addition of each genetic variable.

As a secondary analysis, we compared the results with that of random forest algorithm^89^, to investigate whether this could handle better than logistic regression the multiple predictor variables as well as their potential interactions.

### Metrics of predictive performance

To ensure that models can be utilised across various settings, we chose not to set a specific probability threshold for predicting a “case”. This approach avoids oversimplifying nuanced probability predictions into mere classification and allows the probability decision threshold to be adjusted according to other factors in the process of shared decision making. Therefore, we chose predictive ability metrics that do not rely on a predetermined decision threshold: (1) the area under the ROC curve (AUC ROC)^90^, which evaluates how well the model distinguishes cases from non-cases, (2) the area under the PPV-sensitivity curve^35^ (also known as the precision-recall curve), and (3) the area under the NPV-specificity curve. To accurately estimate out-of-sample predictive performance, we calculated these metrics independently within each of the 10 folds in the SPARK cohort’s cross-validation, thus ensuring that the training and validation data remained distinct, and then computed the average performance across all folds. To obtain PPV-sensitivity and NPV-specificity curves, we computed those curves for each fold in the SPARK dataset, calculated the mean across all folds within 100 sensitivity or specificity bins, and then adjusted LOESS regressions to the results using the GAMLSS package.^87^ Finally, we tested out-of-sample prediction of the model trained on the complete SPARK sample to generalise on SSC and MSSNG samples.

### Cognitive and adaptive scales

Verbal and non-verbal IQ scores were assessed using WISC-IV, DAS-II E-Y, DAS-II S-A, or WASI-I, and Vineland Adaptive Behavior Scales^65^ were assessed in n=1054 participants from the SSC sample. Among these, the motor skills subscale was assessed for a subset of n=160 participants for whom the examiners suspected motor impairments. To assess the correlations among these subscales, quantile regressions using the method of fractional polynomials^91^ were fitted with the GAMLSS package^87^. We evaluated the predictive performance for each genetic and developmental predictor as the variance explained (R^2^), while subtracting that from covariates (sex and first 10 genetic principal components).

### Confidence intervals and statistical tests

Confidence intervals on performance metrics were computed using bootstrap methods with 10,000 iterations. In addition, the statistical significance of improvement in performance when adding new predictors to the model was assessed using the likelihood-ratio test which was previously shown to have superior statistical properties compared to the bootstrap method.^92^ All reported p-values were adjusted for multiple comparisons using the Benjamini–Yekutieli method, and the statistical significance was fixed to p<0.05. Analyses were conducted with R-Studio version 4.3.2 and Scikit-Learn^89^ with Python version 3.11.6.

### Reporting Standards

This study adhered to TRIPOD statement^93^ and the Polygenic Risk Score Reporting Standards (PRS-RS).^94^

## Ethics

This study was approved by the CHU Sainte-Justine Research Centre institutional review board.

## Data

We are grateful to all of the families in SPARK, the SPARK clinical sites and SPARK staff. We appreciate obtaining access to phenotypic and genetic data on SFARI Base.

We thank the families participating in MSSNG as well as the generosity of the donors who support this program. We acknowledge the resources of MSSNG (www.mss.ng), Autism Speaks and The Centre for Applied Genomics at The Hospital for Sick Children, Toronto, Canada.

We are grateful to all of the families at the participating Simons Simplex Collection (SSC) sites, as well as the principal investigators (A. Beaudet, R. Bernier, J. Constantino, E. Cook, E. Fombonne, D. Geschwind, R. Goin-Kochel,

E. Hanson, D. Grice, A. Klin, D. Ledbetter, C. Lord, C. Martin, D. Martin, R. Maxim, J. Miles, O. Ousley, K. Pelphrey, B. Peterson, J. Piggot, C. Saulnier, M. State, W. Stone, J. Sutcliffe, C. Walsh, Z. Warren, E. Wijsman).

This research was enabled by support from Calcul Quebec (http://www.calculquebec.ca) and Compute Canada (http://www.computecanada.ca).

## CONTRIBUTIONS

VRB, SJ and VW designed the study and interpreted the results with significant input from RAIB, LM and SBC.

VRB conducted the analyses. RAIB, AL and GH provided statistical and methodological input. Z.Schmilovich, with the guidance of GAR and SJ; and GH, CP, T.Renne, MJL and Z.Saci, with the guidance of SJ, contributed in preparing the genetic datasets.

JV, AL, JE, AO, XZ and T.Rolland gave input on interpreting the results.

VRB wrote the manuscript with the guidance of SJ and VW, and with inputs from all authors.

## DATA AVAILABILITY

Approved researchers can obtain the SPARK and SSC population datasets described in this study by applying at https://base.sfari.org and to the MSSNG dataset at https://research.mss.ng/.

## REFERENCES

1. Zwaigenbaum, L. & Penner, M. Autism spectrum disorder: advances in diagnosis and evaluation. BMJ 361, k1674 (2018).

2. Szatmari, P. et al. Prospective Longitudinal Studies of Infant Siblings of Children With Autism: Lessons Learned and Future Directions. J. Am. Acad. Child Adolesc. Psychiatry 55, 179–187 (2016).

3. Ozonoff, S. et al. A prospective study of the emergence of early behavioral signs of autism. J. Am. Acad. Child Adolesc. Psychiatry 49, 256–66.e1–2 (2010).

4. Kuo, S. S. et al. Developmental Variability in Autism Across 17 000 Autistic Individuals and 4000 Siblings Without an Autism Diagnosis: Comparisons by Cohort, Intellectual Disability, Genetic Etiology, and Age at Diagnosis. JAMA Pediatr. 176, 915–923 (2022).

5. Ozonoff, S. et al. Diagnostic stability in young children at risk for autism spectrum disorder: a baby siblings research consortium study. J. Child Psychol. Psychiatry 56, 988–998 (2015).

6. Mottron, L., Dawson, M., Soulières, I., Hubert, B. & Burack, J. Enhanced perceptual functioning in autism: an update, and eight principles of autistic perception. J. Autism Dev. Disord. 36, 27–43 (2006).

7. Mottron, L., Gagnon, D. & Courchesne, V. Severity should be distinguished from prototypicality. Autism research: official journal of the International Society for Autism Research vol. 16 1658–1659 (2023).

8. Woolfenden, S., Sarkozy, V., Ridley, G. & Williams, K. A systematic review of the diagnostic stability of Autism Spectrum Disorder. Res. Autism Spectr. Disord. 6, 345–354 (2012).

9. Elias, R. & Lord, C. Diagnostic stability in individuals with autism spectrum disorder: insights from a longitudinal follow-up study. J. Child Psychol. Psychiatry 63, 973– 983 (2022).

10. Pickles, A., McCauley, J. B., Pepa, L. A., Huerta, M. & Lord, C. The adult outcome of children referred for autism: typology and prediction from childhood. J. Child Psychol. Psychiatry 61, 760–767 (2020).

11. O’Nions, E. et al. Autism in England: assessing underdiagnosis in a population-based cohort study of prospectively collected primary care data. The Lancet Regional Health – Europe 29, (2023).

12. Elsabbagh, M. et al. Global prevalence of autism and other pervasive developmental disorders. Autism Res. 5, 160–179 (2012).

13. Lord, C. et al. The Lancet Commission on the future of care and clinical research in autism. Lancet 399, 271–334 (2022).

14. McCauley, J. B., Pickles, A., Huerta, M. & Lord, C. Defining Positive Outcomes in More and Less Cognitively Able Autistic Adults. Autism Res. 13, 1548–1560 (2020).

15. Bowe, A. K., Hourihane, J., Staines, A. & Murray, D. M. The predictive value of the ages and stages questionnaire in late infancy for low average cognitive ability at age 5. Acta Paediatr. 111, 1194–1200 (2022).

16. Månsson, J., Stjernqvist, K., Serenius, F., Ådén, U. & Källén, K. Agreement Between Bayley-III Measurements and WISC-IV Measurements in Typically Developing Children. J. Psychoeduc. Assess. 37, 603–616 (2019).

17. Wolff, J. J. & Piven, J. Predicting Autism in Infancy. J. Am. Acad. Child Adolesc. Psychiatry 60, 958–967 (2021).

18. Pellicano, E. et al. A capabilities approach to understanding and supporting autistic adulthood. Nat Rev Psychol 1, 624–639 (2022).

19. World Health Organization. International Classification of Functioning, Disability, and Health: Children & Youth Version : ICF-CY. (World Health Organization, 2007).

20. Lai, M.-C., Anagnostou, E., Wiznitzer, M., Allison, C. & Baron-Cohen, S. Evidence-based support for autistic people across the lifespan: maximising potential, minimising barriers, and optimising the person–environment fit. Lancet Neurol. 19, 434–451 (2020).

21. Hyman, S. L., Levy, S. E., Myers, S. M. & COUNCIL ON CHILDREN WITH DISABILITIES, SECTION ON DEVELOPMENTAL AND BEHAVIORAL PEDIATRICS. Identification, Evaluation, and Management of Children With Autism Spectrum Disorder. Pediatrics 145, (2020).

22. Baribeau, D. A. et al. Developmental implications of genetic testing for physical indications. Eur. J. Hum. Genet. 30, 1297–1300 (2022).

23. Schaaf, C. P. et al. A framework for an evidence-based gene list relevant to autism spectrum disorder. Nat. Rev. Genet. 21, 367–376 (2020).

24. Kaplanis, J. et al. Evidence for 28 genetic disorders discovered by combining healthcare and research data. Nature 586, 757–762 (2020).

25. Bzdok, D., Varoquaux, G. & Steyerberg, E. W. Prediction, Not Association, Paves the Road to Precision Medicine. JAMA Psychiatry 78, 127–128 (2021).

26. Warrier, V. et al. Genetic correlates of phenotypic heterogeneity in autism. Nat. Genet. 54, 1293–1304 (2022).

27. Zhou, X. et al. Integrating de novo and inherited variants in 42,607 autism cases identifies mutations in new moderate-risk genes. Nat. Genet. 54, 1305–1319 (2022).

28. Douard, E. et al. Effect Sizes of Deletions and Duplications on Autism Risk Across the Genome. Am. J. Psychiatry 178, 87–98 (2021).

29. Bzdok, D., Engemann, D. & Thirion, B. Inference and Prediction Diverge in Biomedicine. Patterns (N Y) 1, 100119 (2020).

30. Rolland, T. et al. Phenotypic effects of genetic variants associated with autism. Nat. Med. 29, 1671–1680 (2023).

31. Sánchez, X. C. et al. Comparing Copy Number Variations in a Danish Case Cohort of Individuals With Psychiatric Disorders. JAMA Psychiatry 79, 59–69 (2022).

32. Davies, R. W. et al. Using common genetic variation to examine phenotypic expression and risk prediction in 22q11.2 deletion syndrome. Nat. Med. 26, 1912– 1918 (2020).

33. Kuchenbaecker, K. B. et al. Evaluation of Polygenic Risk Scores for Breast and Ovarian Cancer Risk Prediction in BRCA1 and BRCA2 Mutation Carriers. J. Natl. Cancer Inst. 109, (2017).

34. Elliott, J. et al. Predictive Accuracy of a Polygenic Risk Score–Enhanced Prediction Model vs a Clinical Risk Score for Coronary Artery Disease. JAMA 323, 636–645 (2020).

35. Inouye, M. et al. Genomic Risk Prediction of Coronary Artery Disease in 480,000 Adults: Implications for Primary Prevention. J. Am. Coll. Cardiol. 72, 1883–1893 (2018).

36. Mavaddat, N. et al. Polygenic Risk Scores for Prediction of Breast Cancer and Breast Cancer Subtypes. Am. J. Hum. Genet. 104, 21–34 (2019).

37. van den Broek, J. J., et al. Personalizing breast cancer screening based on polygenic risk and family history. J. Natl. Cancer Inst. 113, 434–442 (2021).

38. Harrell, F. E., Jr, Califf, R. M., Pryor, D. B., Lee, K. L. & Rosati, R. A. Evaluating the yield of medical tests. JAMA 247, 2543–2546 (1982).

39. Kenny, L. et al. Which terms should be used to describe autism? Perspectives from the UK autism community. Autism 20, 442–462 (2016).

40. Barger, B. D., Campbell, J. M. & McDonough, J. D. Prevalence and onset of regression within autism spectrum disorders: a meta-analytic review. J. Autism Dev. Disord. 43, 817–828 (2013).

41. Karczewski, K. J. et al. The mutational constraint spectrum quantified from variation in 141,456 humans. Nature 581, 434–443 (2020).

42. Dosman, C. F., Andrews, D. & Goulden, K. J. Evidence-based milestone ages as a framework for developmental surveillance. Paediatr. Child Health 17, 561–568 (2012).

43. Savage, J. E. et al. Genome-wide association meta-analysis in 269,867 individuals identifies new genetic and functional links to intelligence. Nat. Genet. 50, 912–919 (2018).

44. Grove, J. et al. Identification of common genetic risk variants for autism spectrum disorder. Nat. Genet. 51, 431–444 (2019).

45. Bölte, S. & Poustka, F. The relation between general cognitive level and adaptive behavior domains in individuals with autism with and without co-morbid mental retardation. Child Psychiatry Hum. Dev. 33, 165–172 (2002).

46. Schatz, J. & Hamdan-Allen, G. Effects of age and IQ on adaptive behavior domains for children with autism. J. Autism Dev. Disord. 25, 51–60 (1995).

47. Dawson, M., Soulières, I., Gernsbacher, M. A. & Mottron, L. The level and nature of autistic intelligence. Psychol. Sci. 18, 657–662 (2007).

48. Lipkin, P. H., Macias, M. M. & COUNCIL ON CHILDREN WITH DISABILITIES, SECTION ON DEVELOPMENTAL AND BEHAVIORAL PEDIATRICS. Promoting Optimal Development: Identifying Infants and Young Children With Developmental Disorders Through Developmental Surveillance and Screening. Pediatrics 145, (2020).

49. Chen, C.-Y. et al. The impact of rare protein coding genetic variation on adult cognitive function. Nat. Genet. 55, 927–938 (2023).

50. Kingdom, R., Beaumont, R. N., Wood, A. R., Weedon, M. N. & Wright, C. F. Genetic modifiers of rare variants in monogenic developmental disorder loci. Nat. Genet. 1– 8 (2024).

51. Wray, N. R. et al. From Basic Science to Clinical Application of Polygenic Risk Scores: A Primer. JAMA Psychiatry 78, 101–109 (2021).

52. Lichtenstein, P. et al. Familial risk and heritability of intellectual disability: a population-based cohort study in Sweden. J. Child Psychol. Psychiatry 63, 1092– 1102 (2022).

53. Plomin, R. & von Stumm, S. The new genetics of intelligence. Nat. Rev. Genet. 19, 148–159 (2018).

54. Yengo, L. et al. A saturated map of common genetic variants associated with human height. Nature 610, 704–712 (2022).

55. Myers, S. et al. Insufficient evidence for ‘autism-specific’ genes. Am. J. Hum. Genet. 106, 587–595 (2020).

56. Satterstrom, F. K. et al. Large-Scale Exome Sequencing Study Implicates Both Developmental and Functional Changes in the Neurobiology of Autism. Cell 180, 568–584.e23 (2020).

57. Arnett, A. B. et al. Developmental Predictors of Cognitive and Adaptive Outcomes in Genetic Subtypes of Autism Spectrum Disorder. Autism Res. 13, 1659–1669 (2020).

58. Ozonoff, S., Li, D., Deprey, L., Hanzel, E. P. & Iosif, A.-M. Reliability of parent recall of symptom onset and timing in autism spectrum disorder. Autism 22, 891–896 (2018).

59. Mitchell, R. E. et al. Strategies to investigate and mitigate collider bias in genetic and Mendelian randomisation studies of disease progression. PLoS Genet. 19, e1010596 (2023).

60. Rødgaard, E.-M., Jensen, K., Miskowiak, K. W. & Mottron, L. Representativeness of autistic samples in studies recruiting through social media. Autism Res. 15, 1447– 1456 (2022).

61. SPARK Consortium. Electronic address: & SPARK Consortium. SPARK: A US Cohort of 50,000 Families to Accelerate Autism Research. Neuron 97, 488–493 (2018).

62. Fischbach, G. D. & Lord, C. The Simons Simplex Collection: a resource for identification of autism genetic risk factors. Neuron 68, 192–195 (2010).

63. C Yuen, R. K., et al. Whole genome sequencing resource identifies 18 new candidate genes for autism spectrum disorder. Nat. Neurosci. 20, 602–611 (2017).

64. Howlin, P., Savage, S., Moss, P., Tempier, A. & Rutter, M. Cognitive and language skills in adults with autism: a 40-year follow-up. J. Child Psychol. Psychiatry 55, 49– 58 (2014).

65. Sparrow, S., Cicchetti, D. & McColl, E. Vineland Adaptive Behavior Scales Interview Edition expanded form manual. (2015).

66. Purcell, S. et al. PLINK: a tool set for whole-genome association and population-based linkage analyses. Am. J. Hum. Genet. 81, 559–575 (2007).

67. Hofmeister, R. J., Ribeiro, D. M., Rubinacci, S. & Delaneau, O. Accurate rare variant phasing of whole-genome and whole-exome sequencing data in the UK Biobank. Nat. Genet. 55, 1243–1249 (2023).

68. Danecek, P. et al. Twelve years of SAMtools and BCFtools. Gigascience 10, (2021).

69. Marees, A. T. et al. A tutorial on conducting genome-wide association studies: Quality control and statistical analysis. Int. J. Methods Psychiatr. Res. 27, e1608 (2018).

70. Manichaikul, A. et al. Robust relationship inference in genome-wide association studies. Bioinformatics 26, 2867–2873 (2010).

71. McCarthy, S. et al. A reference panel of 64,976 haplotypes for genotype imputation. Nat. Genet. 48, 1279–1283 (2016).

72. Pain, O. et al. Evaluation of polygenic prediction methodology within a reference-standardized framework. PLoS Genet. 17, e1009021 (2021).

73. Antaki, D. et al. A phenotypic spectrum of autism is attributable to the combined effects of rare variants, polygenic risk and sex. Nat. Genet. 54, 1284–1292 (2022).

74. Wang, K. et al. PennCNV: an integrated hidden Markov model designed for high-resolution copy number variation detection in whole-genome SNP genotyping data. Genome Res. 17, 1665–1674 (2007).

75. Colella, S. et al. QuantiSNP: an Objective Bayes Hidden-Markov Model to detect and accurately map copy number variation using SNP genotyping data. Nucleic Acids Res. 35, 2013–2025 (2007).

76. Sanders, S. J. et al. Multiple recurrent de novo CNVs, including duplications of the 7q11.23 Williams syndrome region, are strongly associated with autism. Neuron 70, 863–885 (2011).

77. Trost, B. et al. Genomic architecture of autism from comprehensive whole-genome sequence annotation. Cell 185, 4409–4427.e18 (2022).

78. Huguet, G. et al. Genome-wide analysis of gene dosage in 24,092 individuals estimates that 10,000 genes modulate cognitive ability. Mol. Psychiatry 26, 2663– 2676 (2021).

79. Huguet, G. et al. Measuring and Estimating the Effect Sizes of Copy Number Variants on General Intelligence in Community-Based Samples. JAMA Psychiatry 75, 447–457 (2018).

80. Martin, F. J. et al. Ensembl 2023. Nucleic Acids Res. 51, D933–D941 (2023).

81. Samocha, K. E., et al. Regional missense constraint improves variant deleteriousness prediction. bioRxiv 148353 (2017) doi:10.1101/148353.

82. Lord, C., Rutter, M. & Le Couteur, A. Autism Diagnostic Interview-Revised: a revised version of a diagnostic interview for caregivers of individuals with possible pervasive developmental disorders. J. Autism Dev. Disord. 24, 659–685 (1994).

83. Sheldrick, R. C. et al. Establishing New Norms for Developmental Milestones. Pediatrics 144, (2019).

84. Gagnon, D. et al. Bayonet-shaped language development in autism with regression: a retrospective study. Mol. Autism 12, 35 (2021).

85. Poldrack, R. A., Huckins, G. & Varoquaux, G. Establishment of Best Practices for Evidence for Prediction: A Review. JAMA Psychiatry 77, 534–540 (2020).

86. Stone, M. Cross-validatory choice and assessment of statistical predictions. J. R. Stat. Soc. 36, 111–133 (1974).

87. Rigby, R. A. & Stasinopoulos, D. M. Generalized Additive Models for Location, Scale and Shape. J. R. Stat. Soc. Ser. C Appl. Stat. 54, 507–554 (2005).

88. Hujoel, M. L. A., Loh, P.-R., Neale, B. M. & Price, A. L. Incorporating family history of disease improves polygenic risk scores in diverse populations. Cell Genom 2, (2022).

89. Pedregosa, F. et al. Scikit-learn: Machine Learning in Python. J. Mach. Learn. Res. abs/1201.0490, (2011).

90. Bradley, A. P. The use of the area under the ROC curve in the evaluation of machine learning algorithms. Pattern Recognit. 30, 1145–1159 (1997).

91. Royston, P. & Altman, D. Regression using fractional polynomials of continuous covariates: parsimonious parametric modelling. Insur. Math. Econ. 2, 165–166 (1994).

92. Vickers, A. J., Cronin, A. M. & Begg, C. B. One statistical test is sufficient for assessing new predictive markers. BMC Med. Res. Methodol. 11, 13 (2011).

93. Collins, G. S., Reitsma, J. B., Altman, D. G. & Moons, K. G. M. Transparent Reporting of a multivariable prediction model for Individual Prognosis Or Diagnosis (TRIPOD): the TRIPOD Statement. Br. J. Surg. 102, 148–158 (2015).

94. Wand, H. et al. Improving reporting standards for polygenic scores in risk prediction studies. Nature 591, 211–219 (2021).

